# Altered brain activity and functional connectivity after MDMA-assisted therapy for post-traumatic stress disorder

**DOI:** 10.1101/2022.05.25.22275473

**Authors:** S. Parker Singleton, Julie B. Wang, Michael Mithoefer, Colleen Hanlon, Mark S. George, Annie Mithoefer, Oliver Mithoefer, Allison R. Coker, Berra Yazar-Klosinski, Amy Emerson, Rick Doblin, Amy Kuceyeski

## Abstract

3,4-methylenedioxymethamphetamine-assisted therapy (MDMA-AT) for post-traumatic stress disorder (PTSD) has demonstrated promise in multiple clinical trials. MDMA is hypothesized to facilitate the therapeutic process, in part, by decreasing fear response during fear memory processing while increasing extinction learning. The acute administration of MDMA in healthy controls modifies recruitment of brain regions involved in the hyperactive fear response in PTSD such as the amygdala, hippocampus, and insula. However, to date there have been no neuroimaging studies aimed at directly elucidating the neural impact of MDMA-AT in PTSD patients. We analyzed brain activity and connectivity via functional MRI during both rest and autobiographical memory (trauma and neutral) response before and two-months after MDMA-AT in nine veterans and first-responders with chronic PTSD of 6 months or more. We hypothesized that MDMA-AT would increase amygdala-hippocampus resting-state functional connectivity, however we only found evidence of a trend in the left amygdala – left hippocampus (*t* = -2.91, uncorrected p = 0.0225, corrected p = 0.0901). We also found reduced activation contrast (trauma > neutral) after MDMA-AT in the cuneus. Finally, the amount of recovery from PTSD after MDMA-AT correlated with changes in four functional connections during autobiographical memory recall: the left amygdala – left posterior cingulate cortex (PCC), left amygdala – right PCC, left amygdala – left insula, and left isthmus cingulate – left posterior hippocampus. Amygdala – insular functional connectivity is reliably implicated in PTSD and anxiety, and both regions are impacted by MDMA administration. These findings compliment previous research indicating that amygdala, hippocampus, and insula functional connectivity is a potential target of MDMA-AT, and highlights other regions of interest related to memory processes. More research is necessary to determine if these findings are specific to MDMA-AT compared to other types of treatment for PTSD.

This study: NCT02102802 Parent-study: NCT01211405

## 1. INTRODUCTION

Post-traumatic stress disorder (PTSD), which can arise following exposure to a traumatic event or repeated stressful events and is a debilitating social and economic burden on individuals and their families (Davidson 2000; Gradus 2017). PTSD is associated with an increased fear response (VanElzakker, Staples-Bradley, and Shin 2018) and distressing and intrusive re-experiencing of traumatic memories (Ehlers 2010) that often serves as a barrier to the therapeutic process. This may explain why individuals with the most severe PTSD symptoms after trauma are more likely to end up with chronic PTSD durations (Shalev et al. 2019; van der Mei et al. 2020). Lifetime occurrence of PTSD in the general population is around 8%, and, prevalence is significantly higher in military personnel (17.1%) and first responders (10-32%), individuals on the front lines of societal trauma (Hoge et al. 2004; Javidi and Yadollahie 2012). A meta-analysis of trials for military-related PTSD found that cognitive behavioral therapy and prolonged exposure therapy delivered clinically meaningful symptom improvements in 49-70% of patients, however 60-72% of veterans receiving these therapies still retained their PTSD diagnosis (Steenkamp et al. 2015). Adverse outcomes such as increased symptoms and disengagement from treatment cause many current psychological therapies for PTSD to have high dropout rates (Goetter et al. 2015), especially trauma focused therapies (Lewis et al. 2020; Mott et al. 2014).

One approach to developing more effective psychotherapies for PTSD is to administer a drug alongside psychotherapy to aid the therapeutic process (Feduccia et al. 2018). 3,4-methylenedioxymethamphetamine-assisted therapy (MDMA-AT) is hypothesized to reduce the fear response associated with re-experiencing traumatic memories, and therefore may facilitate tolerable processing of traumatic content in patients with PTSD (Mithoefer et al. 2011). Phase 2 and 3 trials have demonstrated promise for MDMA-AT as a viable treatment for PTSD (Mitchell et al. 2021; Mithoefer et al. 2019; Wang et al. 2021; Jerome et al. 2020). MDMA, particularly the *R*-enantiomer, increases pro-social behavior and enhances fear extinction in mice and this effect appears to be mediated by serotonergic mechanisms (Curry et al. 2018; M. B. Young et al. 2017). In healthy humans, acute administration of MDMA has been shown to enhance positive and reduce negative affect during the recollection of autobiographical memories, while preserving vividness and emotional intensity (Carhart-Harris et al. 2014). In another study, MDMA was found to preserve the memory accuracy when administered during both encoding and retrieval phases, while attenuating the recollection of salient details for both positive and negative memories, suggesting that MDMA alters emotional memory representations (Doss et al. 2018). Again in healthy controls, MDMA was found to enhance fear extinction learning/retention rates compared to placebo when administered during extinction training phases (Vizeli et al. 2022; Maples-Keller et al. 2022). These findings suggest that MDMA may aid the therapeutic process, in part, by enabling patient access to emotionally challenging material and facilitating memory reconsolidation/fear extinction processes (Feduccia and Mithoefer 2018).

Functional magnetic resonance imaging (fMRI) measures changes in regional blood oxygenation over time and is thus used as a proxy for fluctuating neuronal activity. In-scanner environments can be absent of stimuli (resting-state fMRI – thought to measure intrinsic brain activity), or tasks or stimuli may be presented to study the regional brain dynamics underlying specific cognitive processes (Glover 2011). In addition to the study of isolated regional activation changes, functional connectivity (FC) can be assessed to infer interaction between two or more brain regions. FC is defined as the statistical relationship (Pearson correlation in the case of the present study) between two brain regions’ activity over time. These tools have been used to study functioning of brain regions and their networks in a wide range of neuropathology and psychiatric disorders (Zhou et al. 2017; Lucassen et al. 2014; Woodward and Cascio 2015).

PTSD patients have shown altered functioning of the precuneus, posterior cingulate cortex (PCC), anterior cingulate cortex (ACC), insula, prefrontal and frontoparietal regions, as well as the hippocampus and amygdala (Sartory et al. 2013; Harnett, Goodman, and Knight 2020; Abdallah et al. 2017; Lazarov et al. 2017; Malivoire et al. 2018; Brown et al. 2014; Pitman et al. 2012; Gilboa et al. 2004; Fonzo et al. 2021; Rabinak et al. 2011; Belleau et al. 2020; Thome et al. 2020; R. A. Lanius et al. 2006; Sripada et al. 2012), suggesting augmented recruitment of brain regions involved in self-referential processing (Cavanna and Trimble 2006), salient autobiographical memory (Svoboda, McKinnon, and Levine 2006; Spreng, Mar, and Kim 2009; Sestieri et al. 2011; Maddock 1999; Spreng and Grady 2010), and fear and emotion (LeDoux 2003).

The specific effects of MDMA-AT on brain function in individuals with PTSD have not been characterized, but several studies suggest the amygdala and hippocampus may play an important role. The amygdala is broadly associated with fear response, and the hippocampus, associated with learning and memory, may provide contextual information necessary for cognitive-affect during memory recall (LeDoux 2003; Harnett, Goodman, and Knight 2020; Pitman et al. 2012). Sripada and colleagues (2012) found combat veterans with PTSD have decreased amygdala-hippocampal resting-state functional connectivity (RSFC) compared to combat veterans without PTSD, which the authors speculate may represent an inability to contextualize affective information in PTSD. Increased amygdala-hippocampal RSFC following stress/trauma exposure has been shown to correlate with recovery from stress or trauma symptoms (Ben-Zion, Keynan, et al. 2020; Vaisvaser et al. 2013; Fan et al. 2015), suggesting a possible adaptive mechanism to threat exposure. In healthy volunteers, the acute administration of MDMA increases amygdala-hippocampal RSFC (Carhart-Harris, 2015). Intranasal oxytocin also increases amygdala-hippocampal RSFC (Fan et al. 2015), and this effect appears to be mediated by serotonin system signaling (Lan et al. 2022) – two neuromodulators that MDMA significantly increases in the extracellular/plasma concentration of, and that play a crucial role in its effects on pro-social behavior and fear extinction (Dumont et al. 2009; Feduccia and Duvauchelle 2008; de la Torre et al. 2004; Green et al. 2003; Thompson et al. 2007; Nardou et al. 2019; Young et al. 2017). Despite evidence that RSFC between the amygdala and hippocampus is implicated in PTSD and that this connection may be modulated by MDMA, no study to date has shown relationships between changes in these regions’ functional connectivity and the therapeutic effects of MDMA-AT.

Herein, we describe results from a study of combat veterans and first-responders undergoing MDMA-AT for PTSD in a randomized, double-blind, dose-response phase 2 clinical trial (Mithoefer et al. 2018). The Clinician-Administered PTSD Scale (CAPS-IV) (an hour-long, semi-structured interview centered around an index trauma) (Blake et al. 1990) was assessed throughout the study to track PTSD severity. Enrolled individuals had moderate-to-severe PTSD with a chronic PTSD duration of 6 months or more. Both resting-state and task-fMRI data, acquired while individuals listened to trauma-related and neutral audio scripts, were collected before and two months after MDMA-AT (follow-up scans were collected after the blind was broken).

Prior to analysis, we hypothesized that MDMA-AT would increase RSFC between the amygdala and hippocampus (Sripada et al. 2012; Carhart-Harris et al. 2015; Ben-Zion, Zeevi, et al. 2020; Vaisvaser et al. 2013; Fan et al. 2015; Lan et al. 2022). We further hypothesized that, at baseline, brain activity would be higher during the trauma-related listening task compared with the neutral listening task in regions associated with autobiographical memory, fear, and emotion, and that this effect would be reduced post-treatment (Sartory et al. 2013). In a final set of analyses, we assessed pre-to-post treatment change in the FC of several regions of interest contained within the limbic, salience, and default mode networks known to be hyperactive in PTSD (Etkin and Wager 2007) during the trauma and neutral autobiographical memory task-fMRI scans. FC changes were then correlated with the pre-to-post treatment recovery in overall PTSD symptomatology – as measured by decreases in CAPS-IV total severity scores.

## 2. METHODS

### 2.1 Trial design

The present study analyzed data from a sub-study (NCT02102802) of a Phase 2 randomized, double-blind, dose-response trial of MDMA-AT in veterans and first responders with severe and chronic PTSD (NCT01211405) (Mithoefer et al. 2018). A detailed study description of the parent study can be found in (Mithoefer et al. 2018), and we summarize the study design in the SI. Here, we provide a description of the MRI-based sub-study design.

Participants in the parent study were able to opt into the MRI-based sub-study after which they provided written informed consent approved by the Medical University of South Carolina Institutional Review Board. They were screened for additional neuroimaging related eligibility criteria and were excluded for any conditions that could render MRI unsafe. A script-driven autobiographical memory paradigm was used to assess brain activity during symptom provocation (Ruth A Lanius et al. 2002; Ruth A. Lanius et al. 2004; Mertens et al. 2022). Following baseline CAPS-IV assessment in the parent study, sub-study participants worked with investigators to create two scripts: one describing a personally traumatic event and one reflecting their typical morning routine at home. Two audio recordings, each six minutes in length, were created from the participant’s reading of each script. Each audio recording was divided into two 3-minute blocks for the task-fMRI. All participants, in all arms, were imaged at baseline, prior to therapy, and again at the follow-up visit two months after their final dosing session. LD (N = 2) and MD (N = 2) participants were additionally imaged after the primary endpoint visit in Stage 1 (one month following their second dosing session), however the small sample sizes prevented any meaningful analysis with these scans. The present analysis focuses on the pre- and post-therapy effects of MDMA-AT on fMRI biomarkers, and thus uses the scans collected at pre-treatment (baseline) and at least 2 months after the largest dose of MDMA (follow-up).

### 2.2 MRI acquisition

At each scanning session, participants underwent MRI on a 32 channel 3T Siemens system. T1 anatomical scans with TR/TE=1900/2.34 ms and 0.9×0.9×1.0 mm voxel size were collected, followed by two identical task fMRI (design described below) (TR/TE = 2200/35ms, 3.0 mm isotropic voxel size, length of each scan = 14:25 min) and one resting state fMRI (TR/TE = 2000/30ms, 3.3×3.3×3.0 mm voxel size, length = 5:00 min).

Participants’ 6-minute trauma and neutral audio scripts were divided into two three-minute trauma and neutral blocks each (See 2.1 Trial design for description of audio recordings). During fMRI, participants were presented with the visual cue “allow” and instructed to allow themselves to experience the scripts as their audio recordings were played for both neutral and trauma blocks. Each task scan had an alternating block design (neutral 1, trauma 1, neutral 2, trauma 2) with an 18 second ‘rest’ period at the start of the scan and between each block, and about a minute of rest at the end of the scan. The precise length of each audio block was 2.95 min.

### 2.3 Image preprocessing

FreeSurfer (Dale, Fischl, and Sereno 1999) was applied to the T1s to create white matter (WM), gray matter (GM) and cerebrospinal fluid (CSF) segmentations. FMRIB Software Library (FSL) (Smith et al. 2004) was used for 1) brain extraction, 2) registration between T1s and fMRI’s (brain-boundary registration, non-linear, full-search), 3) high-pass filtering 4) slice-time and 5) motion correction.

### 2.4 Motion

High-motion frames (defined as > 0.9 mm relative framewise displacement (FD); CONN Toolbox standard parameter (Whitfield-Gabrieli and Nieto-Castanon 2012)) were counted as outlier volumes and scrubbed from functional connectivity and activation analyses. The percentage of scrubbed volumes for resting-state scans ranged from 0 to 6.8% (mean = 2.1%) of total volumes. The percentage of scrubbed volumes for task scans ranged from 0 to 11% (mean = 1.7%) of total volumes. The mean composite FD (Power et al. 2012) for each condition (rest and task) was calculated and compared across time-points using two-sided, paired t-tests. Mean FD during resting-state fMRI scans at baseline and follow-up were 0.12(± 0.06 s.d.) mm and 0.12(± 0.07 s.d.) mm, respectively, and were not significantly different from one another (*t*-statistic = - 0.02; p = 0.99). Mean FD during task fMRI scans at baseline and follow-up were 0.22(± 0.16 s.d.) mm and 0.17(± 0.09 s.d.) mm, respectively, and were not significantly different from one another (*t*-statistic = 1.23, p = 0.25).

### 2.5 Activation analysis: brain response to trauma versus neutral audio listening

FSL’s fMRI Expert Analysis Tool (FEAT) (Woolrich et al. 2004) was used for fitting a general linear model (GLM) to the voxelwise timeseries for each task scan after spatial smoothing using a Gaussian kernel function (6 mm full width at half maximum (FWHM)). For 1^st^-level analysis, models were generated for the neutral block, the trauma block, and a contrast of the two (trauma > neutral). Confound explanatory variables (EVs) included the temporal derivative of each block, 5 nuisance regressors each for WM and CSF signal, outlier volumes (spikes in global signal (> 5 standard deviations) and motion (>0.9 mm FD); CONN Toolbox standard parameters (Whitfield-Gabrieli and Nieto-Castanon 2012)), and 24 motion confounds (Friston et al. 1996). Second-level analysis averaged the models from each of the two task scans performed at each time point. Third-level analyses, using a two-sided, one-sample t-test (FSL randomize; Winkler et al. 2014) identified group-level response for the contrast model (i) at baseline, and (ii) at the two-month follow-up. A final third-level analysis (iii) compared the group-level responses to the contrast model at baseline and follow-up using a two-sample, two-sided, paired t-test (FSL randomize; Winkler et al. 2014). Third-level results were corrected for multiple comparisons using threshold-free cluster enhancement (TFCE; alpha = 0.05) (Smith and Nichols 2009) which identifies significant clusters based on the extent of local support from surrounding voxels.

### 2.6 RSFC analysis

Prior to extraction of RSFC, in addition to the preprocessing steps taken in 2.3, fMRI data were further denoised using an in-house pipeline (https://github.com/kjamison/fmriclean). FMRIs were bandpass filtered and regressed for 24 motion confounds (Friston et al. 1996), 5 nuisance regressors each for WM and CSF, and one for global GM signal. The first five frames (scanner start-up noise) and confound frames (spikes in global signal and motion) were discarded. RSFC (Fisher Z-transformed Pearson correlation values) between the right and left amygdala and right and left hippocampus (Desikan et al. 2006) was calculated for each resting-state scan. For the supplemental RSFC analysis, each hippocampus was further segmented into head and tail portions using FreeSurfer’s hippocampal subregion segmentation tool (Iglesias et al. 2015). Two-tailed, paired *t*-tests were used to compare the 4 RSFC measures before and after MDMA-AT. Baseline to follow-up changes in these 4 measures were also correlated (Pearson’s) with individual level reductions in CAPS-IV using participant’s age and mean FD changes (follow-up – baseline) as covariates of non-interest. All statistical tests were performed at an alpha level of 0.05. P-values were corrected for multiple comparisons using the Benjamini-Hochberg algorithm (Benjamini and Hochberg 1995) where indicated (pFDR).

### 2.7 Task FC analysis

Prior to extraction of task FC, the preprocessed residuals from the task activation analysis (2.5) were further denoised with bandpass filtering and regressed for global GM signal. The first five frames (scanner start-up noise) and confound frames (spikes in global signal (> 5 standard deviations) and motion (>0.9 mm FD); CONN Toolbox standard parameters (Whitfield-Gabrieli and Nieto-Castanon 2012)) were ignored. Functional connectivity (Fisher Z-transformed Pearson correlation values) during each task fMRI scan (task FC) was calculated between 18 regions of interest (ROIs): the right and left hippocampus head, hippocampus tail, amygdala, precuneus, caudal anterior cingulate cortex (ACC), rostral ACC, posterior cingulate cortex (PCC), isthmus cingulate, and insula. All ROIs were extracted from the Disikan-Killiany cortical atlas (see SI Figures 7 and 8 for ROI definitions) (Desikan et al. 2006), and head and tail portions of the hippocampus were created using FreeSurfer’s hippocampal subregion segmentation tool (Iglesias et al. 2015). Two identical task scans were collected at each time point, thus FC values obtained from both scans were averaged to give a single value for each connection per subject. Group-level changes from pre- to post-therapy in the strength of functional connections were assessed using two-tailed, paired *t*-tests. Pearson correlations were calculated between individuals’ changes in functional connection strength and change in CAPS-IV total severity scores (follow-up – baseline) using participant age and mean FD changes between baseline and follow-up as covariates of non-interest. All statistical tests were performed at an alpha level of 0.05. P-values were corrected for multiple comparisons using the Benjamini-Hochberg algorithm (Benjamini and Hochberg 1995) where indicated (pFDR).

## 3. RESULTS

### 3.1 CAPS-IV total severity scores significantly decreased after HD MDMA-AT

Ten participants enrolled in the sub-study, and one withdrew consent after baseline due to anxiety in the MRI scanner, leaving nine participants with MRI data at both time points (6 male, 3 female, aged 41.3; standard deviation (SD) = ± 10.9 years; 8 veterans and 1 first-responder; see SI Table 1 for additional demographic information). All participants had chronic PTSD (mean duration = 84 (± 45) months). One participant’s baseline resting-state fMRI was truncated due to technical issues, leaving eight participants for resting-state analysis and nine for the task fMRI analysis. One participant began the trial with moderate PTSD (CAPS-IV > 39), while the remaining eight presented with severe PTSD (CAPS-IV >59). Mean (SD) CAPS-IV total severity scores of the nine individuals pre- and post-MDMA-AT were 86 (± 16) and 39 (± 25), respectively, representing a significant decrease in PTSD symptom severity between the two time points (Figure 2; N=9, *t* = 6.36, p = 0.00022). The average percent decrease in CAPS was 57 (± 26)%. Results on all participants enrolled in the Phase 2 parent trial have been previously reported (Mithoefer et al. 2018).

### 3.2 Baseline versus two-month follow-up amygdala-hippocampal RSFC

The RSFC was assessed between the amygdala and hippocampus before and after MDMA-AT and the strengths of these connections are illustrated in Figure 3. All connections trended towards increased RSFC after therapy compared to before therapy (using a two-sided paired t-test), with left amygdala to left hippocampus having a significant increase prior to (but not after) corrections for multiple comparisons (N=8, *t* = -2.91, uncorrected p = 0.0225, pFDR = 0.0901). Individual-level pre-to-post-therapy changes between the strength of these functional connections were then correlated (two-sided Pearson’s) with changes in CAPS scores (SI Figure 1). Only one of these correlations (right amygdala to left hippocampus FC versus CAPS) was significant before correction (N = 8; R = -0.820, uncorrected p = 0.0460, pFDR = 0.183).

**Figure 1:**
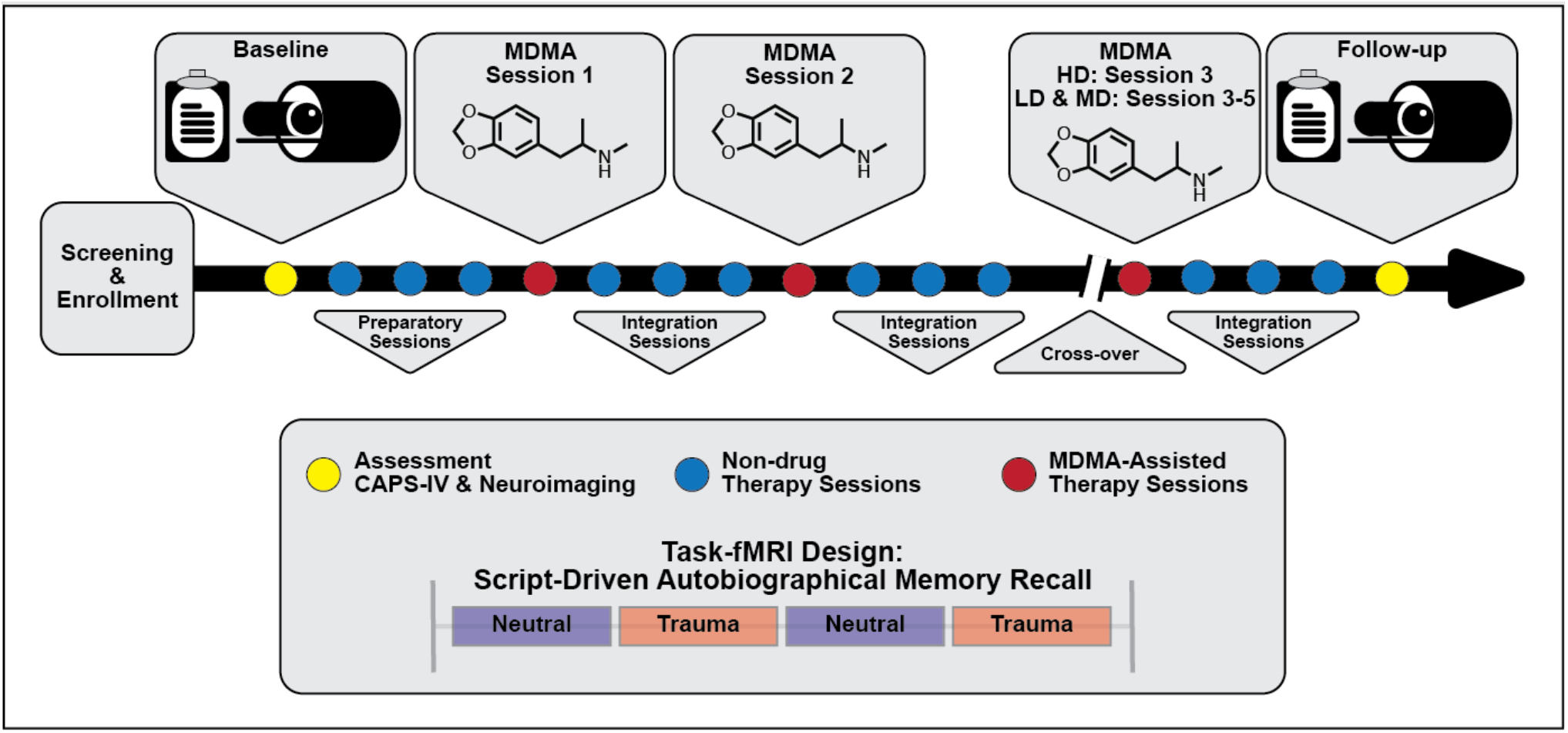
Simplified study design. Subjects were assessed and imaged at the start of the study (baseline). All subjects (low dose (LD; 30 mg MDMA), medium dose (MD; 75 mg MDMA), and high dose (HD; 125 mg MDMA)) underwent three non-drug preparatory therapy sessions prior to their first MDMA dosing session. Each MDMA session was followed by three non-drug integration therapy sessions. After MDMA Session 2 and the subsequent integration sessions, subjects were assessed and the dosing blind was broken. HD subjects completed their final set of drug and non-drug therapy sessions unblinded, and LD/MD subjects crossed over into the HD arm where they completed three sets of drug and non-drug sessions, now with the higher dose and unblinded. All subjects were assessed and underwent MRI approximately two months following their last HD MDMA session. See the Methods section for a full description of study design and scanning protocols.

### 3.4 Brain activation during symptom provocation

A script-driven autobiographical memory paradigm was used to assess brain activity during symptom provocation (Ruth A Lanius et al. 2002; Ruth A. Lanius et al. 2004; Mertens et al. 2022). We compared whole-brain, script-driven activations (trauma > neutral) at baseline and follow-up (Figure 4). Before therapy (baseline), there tended to be larger magnitude activation in response to the trauma script versus the neutral script, as evidenced by the generally positive t-statistics (Figure 4A-D). After correction using threshold free cluster enhancement (TFCE), there was significantly greater activation during the trauma scripts compared to the neutral scripts in four separate areas (see Figure 4 caption for details of each). After therapy, there were smaller magnitude differences between brain activity in response to the two scripts, with no significant clusters (Figure 4E). Finally, we assessed the differences in the contrast before and after therapy (follow-up > baseline). There was generally greater contrast between the trauma and neutral scripts at baseline compared to at follow-up, with one cluster in the bilateral cuneus and lingual gyrus demonstrating significance after correction using TFCE (Figure 4F).

### 3.4 Baseline versus two-month follow-up changes in task FC

We compared the pre- and post-therapy FC strength between 18 brain regions of interest (ROIs) during the task fMRI scans involving neutral and traumatic autobiographical audio recordings (Figure 5A). The ROIs are as follows: the right and left hippocampus head, hippocampus tail, amygdala, precuneus, caudal anterior cingulate cortex (ACC), rostral ACC, posterior cingulate cortex (PCC), isthmus cingulate, and the insula. Only one functional connection was significantly modified at follow-up compared to baseline: the right amygdala to left caudal ACC (N=9; *t*-statistic = 3.04, p = 0.0148). This finding was no longer significant after corrections for multiple comparisons, however (pFDR = 0.9875).

Individual-level pre-to-post therapy changes in these functional connections were then correlated (two-sided, Pearson’s) with the individual-level reductions in CAPS-IV scores (Figure 5B). Most correlations were positive, meaning that larger reductions in connectivity from pre- to post-therapy corresponded to larger improvements in PTSD symptoms. Four correlations between FC and CAPS-IV changes were significant following multiple comparisons correction: the left amygdala and left PCC (N=9; Pearson’s R = 0.951, pFDR = 0.0462), the left amygdala and right PCC (N=9; Pearson’s R = 0.972, pFDR = 0.0197), the left amygdala and left insula (N=9; Pearson’s R = 0.977, pFDR = 0.0197), and the left isthmus cingulate and left hippocampal tail (N=9; Pearson’s R = 0.947, pFDR = 0.0462) (Figure 5C).

### 3.5 Supplemental analyses

Although not the primary focus of our analysis, we also repeated the previous correlations using other secondary outcome measures in place of CAPS-IV total severity scores. Namely, changes between baseline and follow-up in the BDI-II (depression symptoms), the PSQI (sleep quality), the PTGI (perceived growth following trauma), the DES-II (symptoms of dissociation), and the GAF (general psychological function) were used (SI Figure 2). Following correction for multiple comparisons, none of these correlations were significant.

**Figure 2:**
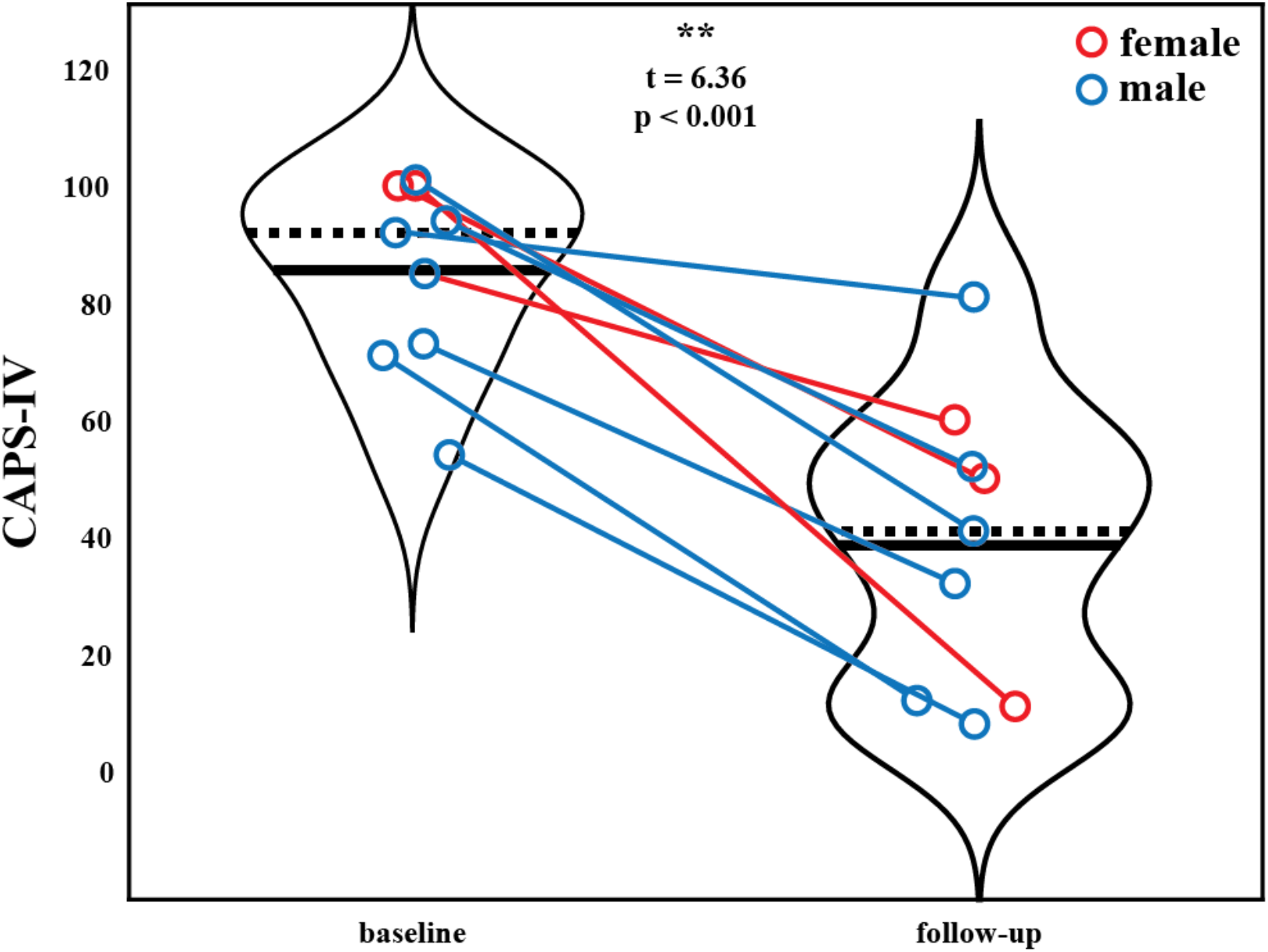
Patient CAPS-IV total severity scores at the baseline (pre-therapy) and two-month follow-up (post-therapy) scanning sessions. Black solid and dashed lines indicate group means and medians, respectively. Blue marker = male. Red marker = female. A significant reduction is PTSD severity was observed after MDMA-AT (baseline > follow-up; N=9, *t* = 6.36, p = 0.00022).

We also replicated our *a priori* analyses of amygdala-hippocampal RSFC using head (anterior) and tail (posterior) sub-regions of the hippocampus (SI Figure 3). The left hippocampal head to left amygdala RSFC was increased at follow-up compared to baseline (N=8; *t* = -2.593, uncorrected p = 0.0358), as was the RSFC between the left hippocampal tail and right amygdala (N=8; *t* = -3.00, uncorrected p = 0.0199). Neither of these effects were significant after corrections for multiple comparisons.

**Figure 3:**
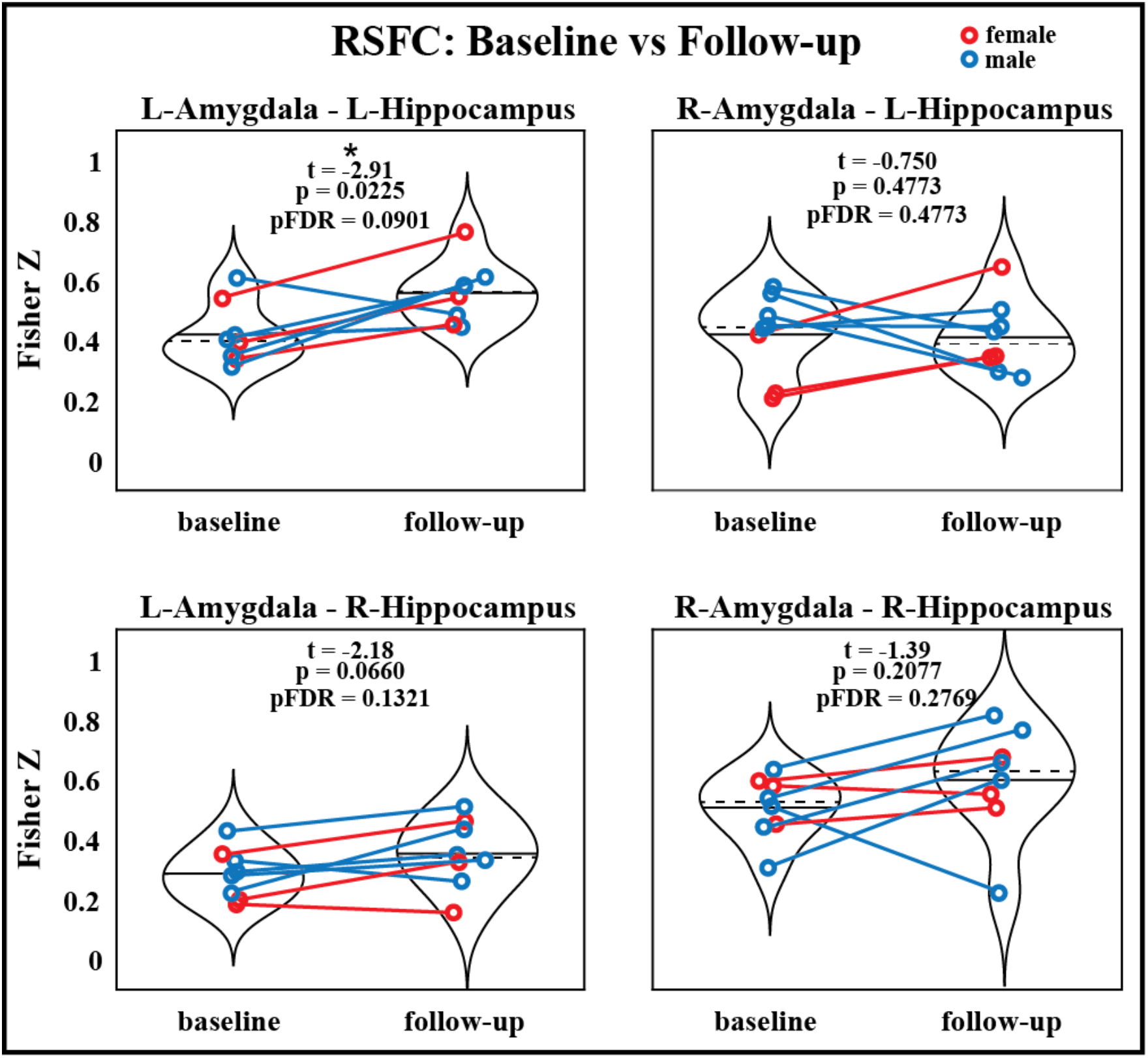
RSFC between the amygdala and hippocampus before and after MDMA-AT. P-values from a two-sided, paired t-test. Blue = male. Red = female. Black solid and dashed lines indicate group means and medians, respectively. (N = 8; t-statistics indicate baseline > follow-up; * uncorrected p < 0.05).

Lastly, we replicate our main functional connectivity analyses without the use of global signal regression and find that these results largely show the same trends, however there is less significance in some cases (SI Figures 4-6). The correlation between the left amygdala and left insula task functional connectivity change and CAPS reductions was significant in both with and without the use of global signal regression (SI Figure 6; N=9; R = 0.971, pFDR = 0.0229).

**Figure 4:**
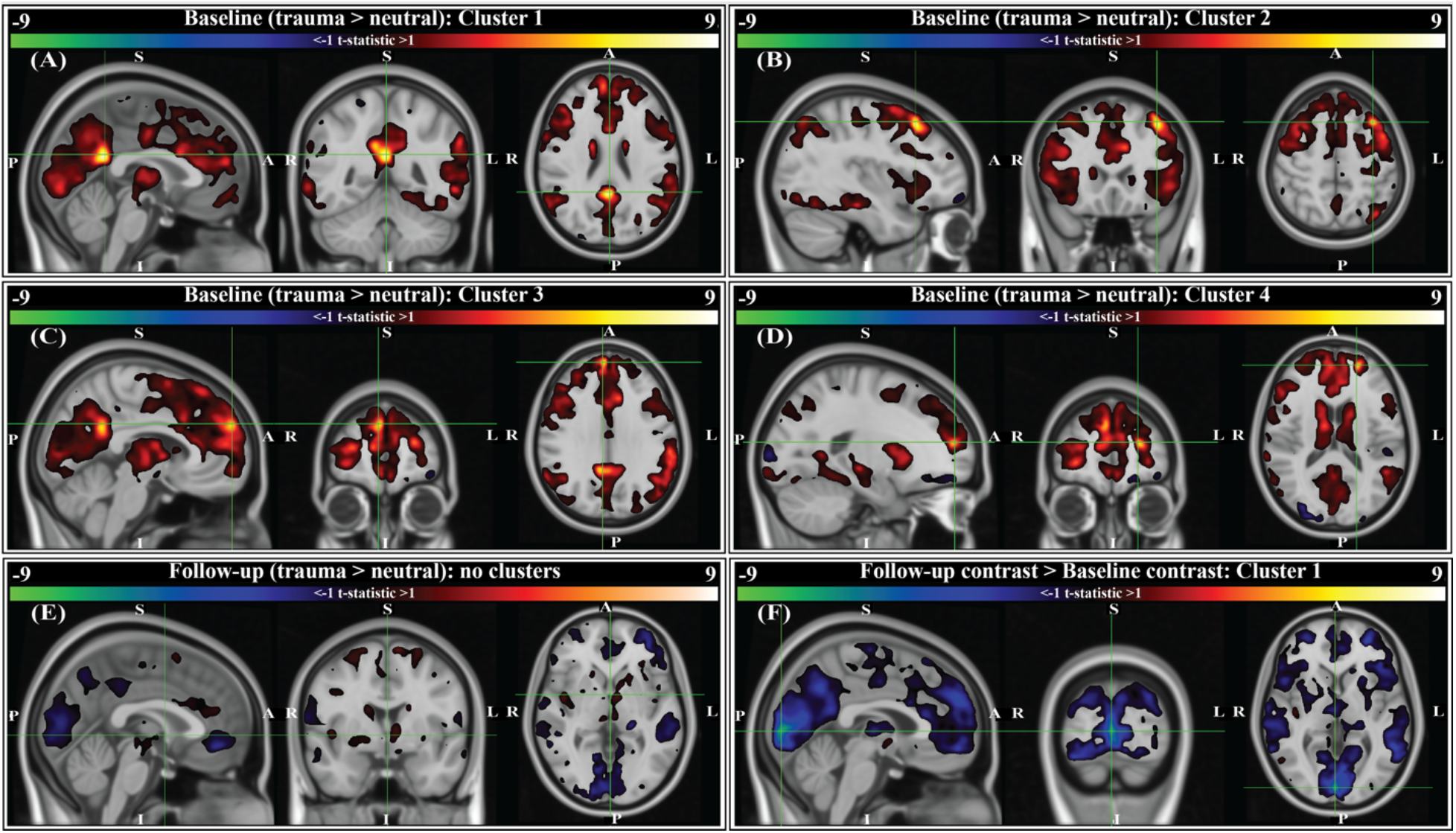
Group-level activation contrasts for trauma versus neutral script listening tasks (N = 9). All panels show t-statistics for corresponding contrasts. Analyses were performed in 3 mm MNI standard space, however results here are interpolated into 1 mm MNI standard space and clipped to only show *t*-statistics greater than +/-1 for visualization purposes. Statistics reported below (voxels, volume, t-statistic, corrected p-value) were calculated from original 3mm results. P-values were corrected using threshold-free cluster enhancement (TFCE; see methods). For panels A-E, positive *t*-statistics indicate greater activation to trauma scripts compared to neutral. For panel F, sign indicates the direction of change in trauma>neutral contrast from baseline (i.e. negative *t*-statistics indicate the contrast between trauma and neutral scripts was decreased at the two-month follow-up compared to baseline). Crosshairs are located on the center of gravity (c.o.g.) of significant clusters. (A) Cluster 1 for the baseline contrast is located primarily in the right and left isthmus cingulate, with some overlap into the right and left precuneus (c.o.g. MNI152 [0, -48, 24]; 6 voxels (162 mm^3^); c.o.g. *t* = 9.61, p(TFCE) = 0.0293). (B) Cluster 2 for the baseline contrast is located in the left caudal middle-frontal gyrus (c.o.g. MNI152 [-36, 21, 51]; 3 voxels (81 mm^3^); c.o.g. *t* = 9.01, p(TFCE) = 0.0234). (C) Cluster 3 for the baseline contrast is located in the right medial prefrontal cortex (c.o.g. MNI152 [6, 57, 30]; 2 voxels (54 mm^3^); c.o.g. *t* = 7.41, p(TFCE) = 0.0488). (D) Cluster 4 for the baseline contrast is located in the left rostral middle frontal gyrus (c.o.g. MNI152 [-21 54 15]; 1 voxel (27 mm^3^); c.o.g. *t* = 9.45, p(TFCE) = 0.0312). (E) There were no significant activation contrasts at the two-month follow-up (crosshairs shown at MNI152 [0, 0, 0]). (F) Comparing the group-level contrasts between time points (follow-up > baseline), there exists one significant cluster located primarily in the right and left cuneus, with some overlap into the right and left lingual gyrus (c.o.g. MNI152 [3, - 90, 3]; 47 voxels (1,269 mm^3^); c.o.g *t* = -9.31, p(TFCE) = 0.0391).

## DISCUSSION

We report signatures of brain response during rest and audio listening task in eight veterans and one first-responder with clinically diagnosed chronic and severe PTSD before and two-months after MDMA-assisted therapy. We found a significant reduction in CAPS-IV total severity scores after therapy (Figure 2), indicating our sub-study participants mirrored the results from the parent study (Mithoefer et al. 2018). We found a trend suggesting that RSFC between the amygdala and hippocampus was strengthened post-therapy, particularly in the left hemisphere (Figure 3). Prior work suggests that modulation of amygdalae-hippocampal RSFC may be an important component of MDMA-AT for PTSD (Sripada et al. 2012; Ben-Zion, Keynan, et al. 2020; Vaisvaser et al. 2013; Fan et al. 2015; Carhart-Harris et al. 2015; Lan et al. 2022), thus investigating this connection in future studies is warranted. We also found participants had increased activation in areas involved with self-referential processing and autobiographical memory while listening to traumatic versus neutral memory narrations pre-therapy (Figure 4A-D), and that no significant contrast existed after MDMA-AT (Figure 4E). Comparing trauma versus neutral contrasts between baseline and follow-up revealed a significant decrease in cuneus contrast after MDMA-AT (Figure 4F). Finally, the pre- to post-therapy reductions in FC during the script listening task between the left amygdala and right PCC, left PCC, and left insula, as well as FC between the left isthmus cingulate and left hippocampal tail strongly and significantly correlated with PTSD symptom improvement (Figure 5C). These results begin to shed light on the neurological mechanisms that may drive MDMA-AT for PTSD.

**Figure 5:**
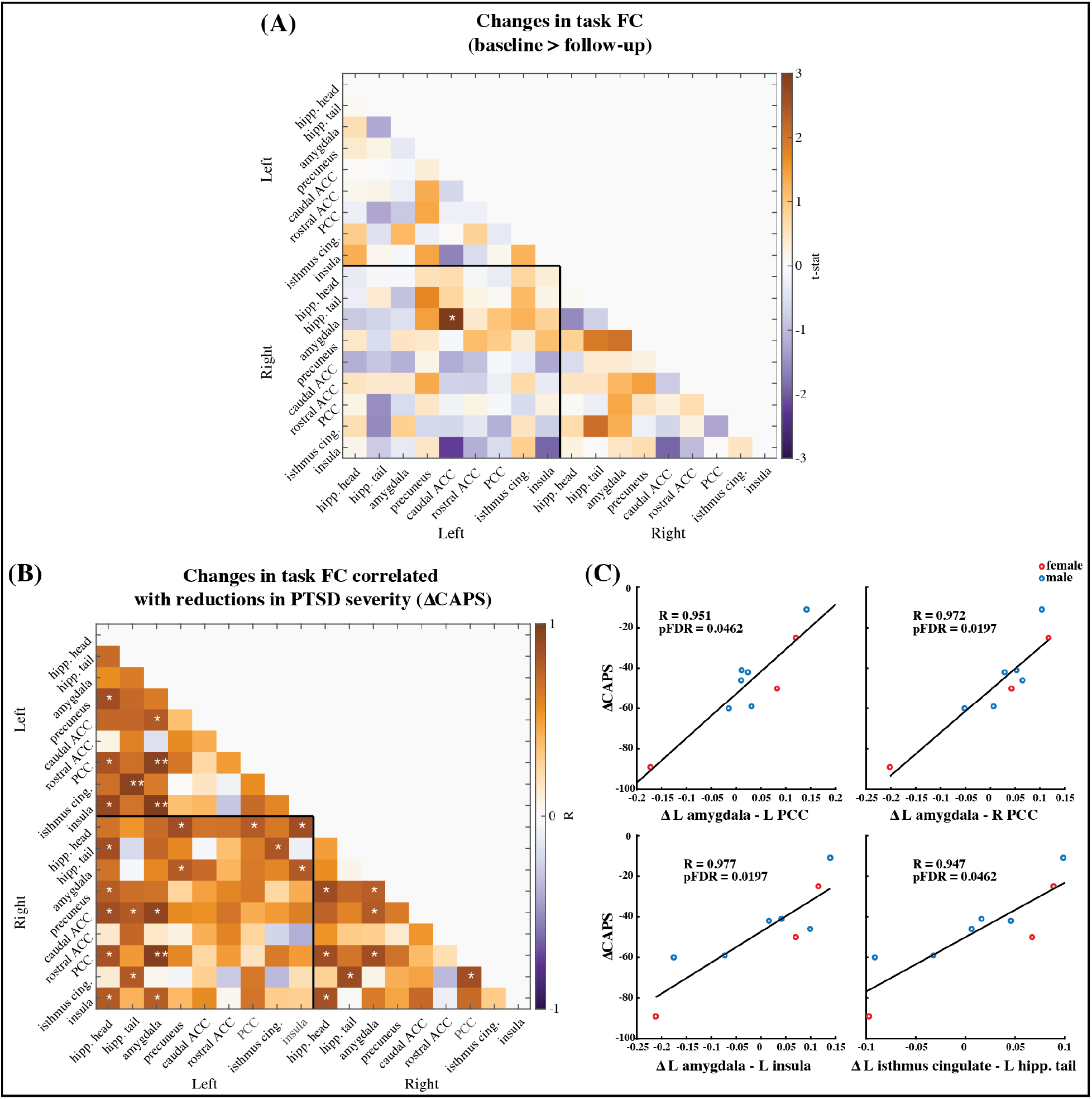
(A) Paired t-statistics shown for differences in functional connectivity between all brain regions of interest during the task fMRI scan involving neutral and trauma memory audio listening (N = 9; baseline > follow-up; * two-sided uncorrected p < 0.05; ** pFDR < 0.05, corrected). (B) Pearson correlation values between changes in ROI functional connectivity and reduction in CAPS scores. Changes were calculated as follow-up values minus baseline values. (N=9; * uncorrected p < 0.05; ** pFDR < 0.05) Age and mean FD difference between baseline and follow-up were included as covariates of non-interest. (C) Scatter plots of the three correlations that remained significant after corrections for multiple comparisons (i.e. pFDR < 0.05). Red marker = female. Blue marker = male.

Previous work quantifying functional connectivity in PTSD and in stress exposure (Sripada et al. 2012; Ben-Zion, Keynan, et al. 2020; Vaisvaser et al. 2013; Fan et al. 2015) and acute MDMA administration in controls (Carhart-Harris et al. 2015) suggests one mechanism of MDMA-AT may be to increase pathologically lowered amygdala-hippocampal RSFC (Feduccia and Mithoefer 2018). The amygdala is associated with fear expression, threat recognition, and heightened response to emotional memories and is often dysregulated in patients with PTSD (LeDoux 2003; Pitman et al. 2012; Liberzon et al. 1999; Etkin and Wager 2007; Bremner et al. 2005; Harnett, Goodman, and Knight 2020). The hippocampus also plays a central role in PTSD as it is thought to provide contextual information important for cognitive-affect during memory recollection (Harnett, Goodman, and Knight 2020; Pitman et al. 2012). Sripada and colleagues (2012) found combat veterans with PTSD had reduced amygdala-hippocampal RSFC compared to combat-exposed controls, leading them to speculate that this may relate to an inability to contextualize affective information in PTSD. Carhart-Harris and colleagues (2015) demonstrated that amygdala-hippocampal RSFC is increased acutely in MDMA administration compared to placebo and this increase occurred in a manner that correlated with the drug’s subjective effects at a near-significant level, leading these researchers to propose that this functional connection was a primary target of MDMA-AT. Increased amygdala-hippocampal RSFC has also been linked to intranasal oxytocin administration after stress exposure (Fan et al. 2015) and this effect was mediated by serotonin signaling (Lan et al. 2022) – two neuromodulators that play a significant role in the pro-social and fear extinction effects of MDMA (Dumont et al. 2009; de la Torre et al. 2004; Feduccia and Duvauchelle 2008; Green et al. 2003; Thompson et al. 2007; Nardou et al. 2019; Young et al. 2017). Prior to our analysis (although after the study was designed and the data collected), we hypothesized that the RSFC between the amygdala and hippocampus would be higher after MDMA-AT compared to pre-therapy levels. Only the RSFC between the left amygdala and left hippocampus was significantly increased (Figure 3) however, this finding no longer met thresholds for significance after multiple comparisons correction (pFDR = 0.09). We also found that the amount of increased right amygdala – left hippocampal RSFC after MDMA-AT positively correlated with PTSD symptom improvement at a near-significant level (SI Figure 1; R = -0.820, uncorrected p = 0.046, pFDR = 0.183). Our current findings, though inconclusive, are suggestive of a trend towards moderate increases in amygdala – hippocampal RSFC two-months after MDMA-AT. It is possible that more significant changes would have been observed with a larger sample size, longer resting-state scans, or imaging performed closer to MDMA administration. These findings justify the continued investigation of amygdala-hippocampal RSFC in the therapeutic mechanisms of MDMA-AT in future studies.

We next sought to study brain response during autobiographical memory listening to draw additional conclusions about MDMA-AT’s effects in individuals with PTSD. Before therapy, participants had larger activation in four areas during an individualized trauma script listening task compared to neutral script listening: the right and left isthmus cingulate and precuneus, the left caudal middle frontal gyrus, the right medial prefrontal cortex, and the left rostral middle frontal gyrus (Figure 4A-D). These regions are broadly involved in self-processing operations (e.g. first-person perspective taking), episodic memory retrieval, visual-spatial imagery, autobiographical memory recollection, and are included in or interact with the default mode network (Cavanna and Trimble 2006; Cauda et al. 2010; Spreng, Mar, and Kim 2009; Spreng and Grady 2010; Kalenzaga et al. 2015; Demblon, Bahri, and D’Argembeau 2016; Raichle 2015). The retrosplenial cortex – located within the isthmus cingulate - is also found to be consistently activated by emotionally salient stimuli, and has been proposed to play a role in the interaction between emotion and memory (Maddock 1999). We conjecture that increased activation in these regions during traumatic compared to neutral audio listening (Figure 4A-D) could be related to an increased intensity of the recollection or re-experiencing of traumatic memories compared to neutral ones for patients before therapy. At 2-month follow-up to MDMA-AT, there was no significant difference in the trauma vs neutral script activation (Figure 4E). The longitudinal comparison of these two time points indicated that the contrast between trauma and neutral was larger at baseline, particularly in a significant cluster in the right and left cuneus/lingual gyrus (Figure 4F). Cuneus activity during autobiographical memory tasks often coincides with activity in the frontal regions highlighted by the baseline contrast, and has been found to correlate with memory recall accuracy (Spreng and Grady 2010; Demblon, Bahri, and D’Argembeau 2016; Kalenzaga et al. 2015). Cuneus activity is thought to enhance the visual imagery of autobiographical memory recollection (Cabeza and St Jacques 2007), therefore decreased contrast in this area at follow-up suggests that intensity of visual imagery contrast between trauma and neutral memories may be decreased after MDMA-AT. Larger studies may allow more statistical power to identify additional longitudinal differences. Other longitudinal studies of individuals with PTSD have found that decreases in precuneus, isthmus cingulate, and middle frontal gyrus activation during symptom provocation is correlated with reductions in PTSD symptom severity (Garrett et al. 2019; Ke et al. 2016).

PTSD is often associated with hyperactivity in the amygdala (Pitman et al. 2012); the acute administration of MDMA in healthy volunteers decreases blood flow to the amygdala during rest (Carhart-Harris et al. 2015) and attenuates its response to angry faces (Bedi et al. 2009). We had hypothesized that we would observe hyperactivity of the amygdala to trauma versus neutral scripts at baseline and that MDMA-AT would attenuate this response, however we observed neither. It is important to note inconsistencies in the literature here. Amygdala hyperactivity in PTSD is not always observed, possibly due to differences in subtypes, sex, cultural representations, or choice of paradigm (van Huijstee and Vermetten 2018; R. A. Lanius et al. 2001; Ruth A Lanius et al. 2002; Helpman et al. 2021; Chiao et al. 2008; Liddell and Jobson 2016). Additionally, while MDMA did suppress amygdala activity during rest and in response to angry faces as previously mentioned, there was no observed impact on its response to autobiographical memories (Carhart-Harris et al. 2014). While activation-based analyses deserve continued attention in future studies to rectify these inconsistencies, functional connectivity is a complimentary approach we can use to extract additional information from fMRI.

Functional connectivity analyses in individuals with PTSD have revealed aberrant connectivity between several regions within default mode, limbic, and salience networks, and more broadly, regions involved in emotional and self-referential processing (Abdallah et al. 2017; Lazarov et al. 2017; Malivoire et al. 2018; Brown et al. 2014; Gilboa et al. 2004; Fonzo et al. 2021; Rabinak et al. 2011; Belleau et al. 2020; Thome et al. 2020; Ruth A. Lanius et al. 2004; Sripada et al. 2012), and, further, the administration of MDMA in healthy volunteers has been shown to disrupt the functional integrity of these networks (Walpola et al. 2017; Carhart-Harris et al. 2015; Müller et al. 2021; Dipasquale et al. 2019; Avram et al. 2022). Here, we measured functional connectivity during script-driven autobiographical memory recall between the right and left hippocampus head, hippocampus tail, amygdala, precuneus, caudal ACC, rostral ACC, PCC, isthmus cingulate, and the insula. Our ROIs were defined and labeled based on the Desikan-Killiany (DK) brain atlas (SI Figures 7 and 8) (Desikan et al. 2006). We chose to segment the hippocampus into anterior (head) and posterior (tail) ROIs based on recent work showing that the two portions’ FC are differentially effected by PTSD (Malivoire et al. 2018). We assessed group-level changes in the strength of these functional connections and found no significant differences between baseline and follow-up after corrections for multiple comparisons (Figure 5A). However, we did find that greater recovery (larger decreases in CAPS-IV at follow-up) was associated with reductions in FC between the left amygdala and the right and left PCC, as well as the left insula (Figure 5C). The acute effects of MDMA in healthy volunteers has been shown to decrease the FC of the PCC (Müller et al. 2021; Dipasquale et al. 2019) and insula (Walpola et al. 2017), and alter amygdala and hippocampus FC (Carhart-Harris et al. 2015), highlighting the potential relevance of our current findings. Amygdala to posterior and mid-cingulate cortex FC has been shown to be associated with PTSD severity at different stages of disease progression, although differing patient populations and assessment time-lines lead to conflicting results (Belleau et al. 2020; R. A. Lanius et al. 2010; Bluhm et al. 2009; Cisler et al. 2016; Shou et al. 2017). One finding in healthy adults shows increased amygdala - PCC FC following the acute exposure to stress (Veer et al. 2011), thus the association between recovery and reduced amygdala – PCC task FC at follow-up possibly relates to reduced stress response to trauma memories (although the finding by Veer and colleagues is more posterior to the ROI used here). Amygdala and insula RSFC is increased in PTSD (Sripada et al. 2012; Rabinak et al. 2011) (except in one study which finds the opposite (Fonzo et al. 2021)), and reduced amygdala-insula FC during negative image reappraisal is associated with larger improvements in PTSD symptoms (Cisler et al. 2016). The strength of left amygdala-insula FC also positively correlates with the amount of acute anxiety measured in participants just before scanning (Baur et al. 2013). Attenuated functional connectivity of these two regions at follow-up in the present study possibly suggests a decreased intensity of recalled events, less ‘re-experiencing’, or reduced anxiety during the script-driven memory recall (Etkin and Wager 2007). Lastly, we found that reductions in CAPS-IV at follow-up were associated with reduced FC between the left isthmus cingulate and left hippocampal tail (Figure 5C). The isthmus cingulate labeled here consists of the most posterior potions of the PCC (SI Figure 7). Increased FC between these two regions has previously been reported in PTSD patients compared to trauma-exposed health controls (Malivoire et al. 2018; Lazarov et al. 2017), and the present finding is possibly indicative of changes in memory contextualization and reduced threat sensitivity at two-month follow-up to MDMA-AT compared to baseline (Bluhm et al. 2009).

PTSD is characterized by decreased fear extinction in response to trauma-related stimuli. One possible mechanism through which MDMA-AT operates is enhanced reconsolidation and/or fear extinction processes (Feduccia and Mithoefer 2018). Several studies with MDMA implicate reconsolidation or fear extinction processes, and while it is currently unclear whether MDMA acts on only one or both, it is important to note that the two interact (Suzuki et al. 2004). Rodent models have demonstrated that the administration of MDMA prior to extinction learning enhances extinction retention (tested 48 hours after learning) and this effect is blocked by acute and chronic treatment with a serotonin transporter inhibitor (M. B. Young et al. 2015; Young 2017).

Hake et al. (2019) found that MDMA administered during extinction learning phases did not enhance fear extinction memory, while MDMA administration during reconsolidation phases resulted in prolonged reductions in conditioned fear. In addition, MDMA administered prior to trauma-cue exposure (reconsolidation phase) in rodents resulted in reduced stress-related behavioral responses 7 days later (Arluk et al. 2022). Two recent trials in healthy humans found that MDMA (100 mg and 125 mg, respectively) administered prior to extinction learning resulted in improved extinction learning at extinction recall phases (48hr and 24hr later, respectively) compared to the placebo group (Maples-Keller et al. 2022; Vizeli et al. 2022). Doss and colleagues (2018) found that 1 mg/kg of MDMA in healthy humans attenuated the encoding and retrieval of salient details from positive and negative stimuli (but not neutral stimuli), suggesting an ability for MDMA to alter emotional memory representation.

Interestingly, a fMRI study in healthy humans found decreased activation in the precuneus/PCC during fear extinction learning (Ridderbusch et al. 2021), regions highlighted by our present study and others in PTSD (Garrett et al. 2019; Ke et al. 2016).

## LIMITATIONS

The small sample size of the present study and the lack of a control population (e.g. trauma-exposed healthy controls) may decrease the generalizability of these findings. The trial design was placebo-controlled for dose-response (low, medium, and high), however, the follow-up scans used in this study were after the breaking of the blind and dose cross-over (low/medium to high) had occurred. For neuroimaging studies, comparisons with control populations are helpful for contextualizing longitudinal changes in brain response and provide information about whether changes in patient populations represent an abnormal response being restored to normality or a compensatory mechanism. In addition, multi-point imaging of healthy control or non-treatment (placebo) groups allow for the quantification of test-retest variability. Lastly, had we imaged a cohort that received therapy without MDMA, we would have been able to assess longitudinal brain changes that were unique to or enhanced by MDMA.

Here it must be discussed that PTSD is a disorder exhibiting at least two major sub-types (dissociative and non-dissociative) with characteristically opposing phenomenological and physiological responses to symptom provocation, which may explain inconsistencies in the PTSD neurobiology literature (van Huijstee and Vermetten 2018). PTSD sub-type information was not collected in the present study. In addition to sub-type heterogeneity, males and females may also differ in their adaptive neural responses to trauma (Helpman et al. 2021). Limited by our sample size, we did not investigate differences between males and females in this study.

The accepted standard for assessing PTSD severity is the Clinician-Administered PTSD Scale (Blake et al. 1990). Specifically, CAPS-IV was used in this study. CAPS-IV involves an hour-long semi-structured interview with a clinician and, though comprehensive, faces limitations. In their baseline CAPS-IV assessment, and subsequently thereafter, patients were asked to refer to an index trauma that was measured throughout the study. This may present an issue in accurately assessing global PTSD severity if an adjacent or un-related trauma surfaces during therapy and becomes the prominent driver of their symptoms. These issues, combined with difficulty in blinding and expectancy effects, present additional challenges in accurately mapping fMRI metrics to clinical outcomes.

The task design used in this study examined differences in brain response to personalized audio scripts generated from narrations of traumatic and neutral memories. Many different stimuli have been used in fMRI studies of PTSD (R. A. Lanius et al. 2006; Sartory et al. 2013; Patel et al. 2012), each providing its own unique advantages and disadvantages. Our present design optimizes personal relevance of the stimuli; however, this has the consequence of presenting each subject with a different set of stimuli, whereby brain responses within each block are not time-locked across participants. Also, it has previously been shown that PTSD survivors take longer to retrieve unrelated autobiographical information when listening to taped imagery scripts of their traumatic memories (Kleim, Wallott, and Ehlers 2008). This suggests the possibility that those with the most severe PTSD will take the longest to cognitively transition to the neutral block from the trauma block. If this is true, then there would perhaps be an inverse-”U” relationship between PTSD severity and contrast between the trauma and neutral conditions, if the blocks are not spaced far enough apart to allow adequate time for patients to return to a baseline level of cognitive functioning. Additionally, because our repeated task fMRI scans were identical (rather than counterbalanced for condition), there could be primacy effects in the neutral condition (which was always first) and/or fatigue effects in the trauma condition (which was always last).

We did not aim to characterize lateralization in our findings, and though most of our significant FC results were found in the left amygdala, we did not test for statistical interaction effects between hemispheres. While lateralization of the amygdala remains debated, it has been suggested by early work that the left amygdala is more strongly related to conscious (versus unconscious) perception and emotional regulation (Morris, Öhman, and Dolan 1998).

Finally, the pre-specified aim of this study was to estimate longitudinal (baseline to 2-months after final MDMA session) changes in ROI response to traumatic audio scripts. Between the start of data collection and analysis, new literature emerged (Carhart-Harris et al. 2015; Lan et al. 2022) implicating amygdala-hippocampus RSFC as a potential target of MDMA-AT, compelling us to expand our analysis beyond the pre-specified aims. Functional connectivity estimates from shorter scans (e.g. five minutes in the case of our resting-state data) can have lower reliability (Birn et al. 2013) and therefore the trends in increased RSFC between amygdala and hippocampal regions reported here should be considered preliminary.

## CONCLUSION

We report functional brain changes associated with MDMA-AT in veterans and first responders with moderate-to-severe and chronic PTSD. We had hypothesized that MDMA-AT may act through strengthening the RSFC between the amygdala and hippocampus, a connection which is weaker in PTSD populations (Sripada et al. 2012) and increased acutely by MDMA in healthy volunteers (Carhart-Harris et al. 2015). The trends found here are suggestive of such in the left amygdala – left hippocampus, however larger studies are needed. We also provide preliminary evidence that MDMA-AT alters brain response during symptom provocation in regions associated with fear response, anxiety, self-referential processing, and salient autobiographical memory, and are commonly found to be hyperactive in PTSD patients (Sartory et al. 2013; Patel et al. 2012). Finally, the reduction of several functional connections during autobiographical memory audio co-varied with symptom reduction in PTSD. Of these connections, the left amygdala – left insula is perhaps the most interesting, due to the role of amygdala-insular FC in anxiety and PTSD symptomatology (Baur et al. 2013; Cisler et al. 2016; Sripada et al. 2012; Rabinak et al. 2011). More research is necessary to confirm these results and to disentangle effects specific to MDMA and its combination with psychotherapy.

## Data Availability

All data in the present study will be made available upon reasonable request to the authors following peer-review and publication.

## Acknowledgements

The authors would like to thank the candidates who were willing to be screened for eligibility; the participants in the trial who contributed their data; S. Sadler for her dedication as the Study Coordinator; S. Braswell for serving as night attendant, M. Wagner and J. Wymer for Independent Rater assessments. The authors would also like to thank: J. Holland who supported this trial as a Medical Monitor; L. Jerome for her numerous and varied contributions to this Clinical Development Program since prior to its inception with global systematic literature reviews and medical coding; C. Hennigan for data management; R. Matthews and B. Shechet for clinical operations and monitoring; J. Sonstroem and A. Seltzer for randomization support and system programming; E. Sola, Y. Gelfand, and B. Cohen for conducting adherence ratings to facilitate standardization of therapy; I. Gorman for development of adherence ratings; A. Wilens for supporting video recording. The authors would like to thank Dr. Edmund Higgins for catalyzing the first discussions about acquiring imaging data in the MAPS trial.

