## Supplementary material for "Altered brain activity and functional connectivity after MDMA-assisted therapy for post-traumatic stress disorder": SI

Trial Design:

Participants were recruited and screened between November 10, 2010 and January 29, 2015. Veterans and first responders with severe PTSD as measured by a Clinician-Administered PTSD Scale (CAPS-IV) (Blake et al. 1990) total severity score of 50 or more were enrolled in the study and received three 90-minutes preparatory therapy sessions. In Stage 1, participants were randomly assigned to three groups (1:1:2) that received blinded 30, 75 or 125 mg MDMA HCl (followed by a supplemental half-dose unless withheld or declined) with therapy in two 8-hour dosing sessions approximately one month (3-5 weeks) apart. Each of the dosing sessions was followed by three non-drug 90-minute follow-up integration sessions, the first occurring the morning after dosing and the remaining two approximately one week apart. Psychological assessments were collected at baseline and the primary endpoint which was one month after the second dosing session. In Stage 2 of the study, the blind was broken and participants originally in the HD (125 mg) group participated in a final open-label session at the same dose, while participants originally in the LD (30 mg) or MD (75 mg) groups participated in three HD (100 – 125 mg) open-label sessions. Each dosing session in Stage 2 was followed by three non-drug 90-minute integrative sessions with the same spacing as Stage 1. A detailed diagram of Stage 1 and 2 is provided on the last page of the Supplementary Information. Participants were required to taper and abstain from psychotropic medications during study participation except for sedative hypnotics or anxiolytics used as needed between MDMA sessions. Psychological assessments were collected approximately 2 months and 12 months after the final dosing session in Stage 2. Psychological assessment included the clinician-administered measures CAPS-IV and Global Assessment of Functioning (GAF; general psychological function; Guze 1995), and self-reported measures including Beck Depression Inventory-II (BDI-II; depression symptoms; Beck et al.

1996), Pittsburgh Sleep Quality Index (PSQI; sleep quality; Buysse et al. 1989), Post-Traumatic Growth Inventory (PTGI; perceived growth following trauma; Tedeschi and Calhoun 1996), and the Dissociative Experiences Scale II (DES-II; symptoms of dissociation; Carlson and Putnam 1993).

| | Mean (SD / %) |
|---|---|
| Mean age, years | 43.1 (10.9) |
| Mean BMI | 29.4 (3.4) |
| Mean duration of PTSD, months | 84 (45) |
| Sex | |
| Male | 6 (67%) |
| Female | 3 (33%) |
| Ethnicity | |
| Caucasian | 8 (89%) |
| Native American | 1 (11%) |
| Occupation associated with trauma | |
| Military | 8 (89%) |
| Police officer | 1 (11%) |
| Pre-study therapy | |
| Eye movement desensitization reprocessing | 1 (11%) |
| Group psychotherapy | 1 (11%) |
| Cognitive processing therapy | 1 (11%) |
| Cognitive behavioral therapy | 8 (89%) |
| Psychodynamic therapy | 3 (33%) |
| None | 1 (11%) |
| Pre-study psychiatric medications | |
| Antidepressants | 9 (100%) |
| Anxiolytics | 7 (78%) |
| Antipsychotics | 2 (22%) |
| Hypnotics and sedatives | 7 (78%) |
| Stimulants | 5 (56%) |
| Psychiatric comorbid disorders | |
| Major depression | 8 (89%) |
| Panic disorder | 5 (56%) |
| Generalized anxiety disorder | 2 (22%) |
| Lifetime C-SSRS | |
| Positive ideation | 5 (56%) |
| Serious ideation | 2 (22%) |
| Positive behavior | 3 (33%) |

SI Table 1: N=9. Demographic information recorded at baseline. BMI=body-mass index. PTSD=post-traumatic stress disorder. C-SSRS=Columbia-Suicide Severity Rating Scale.

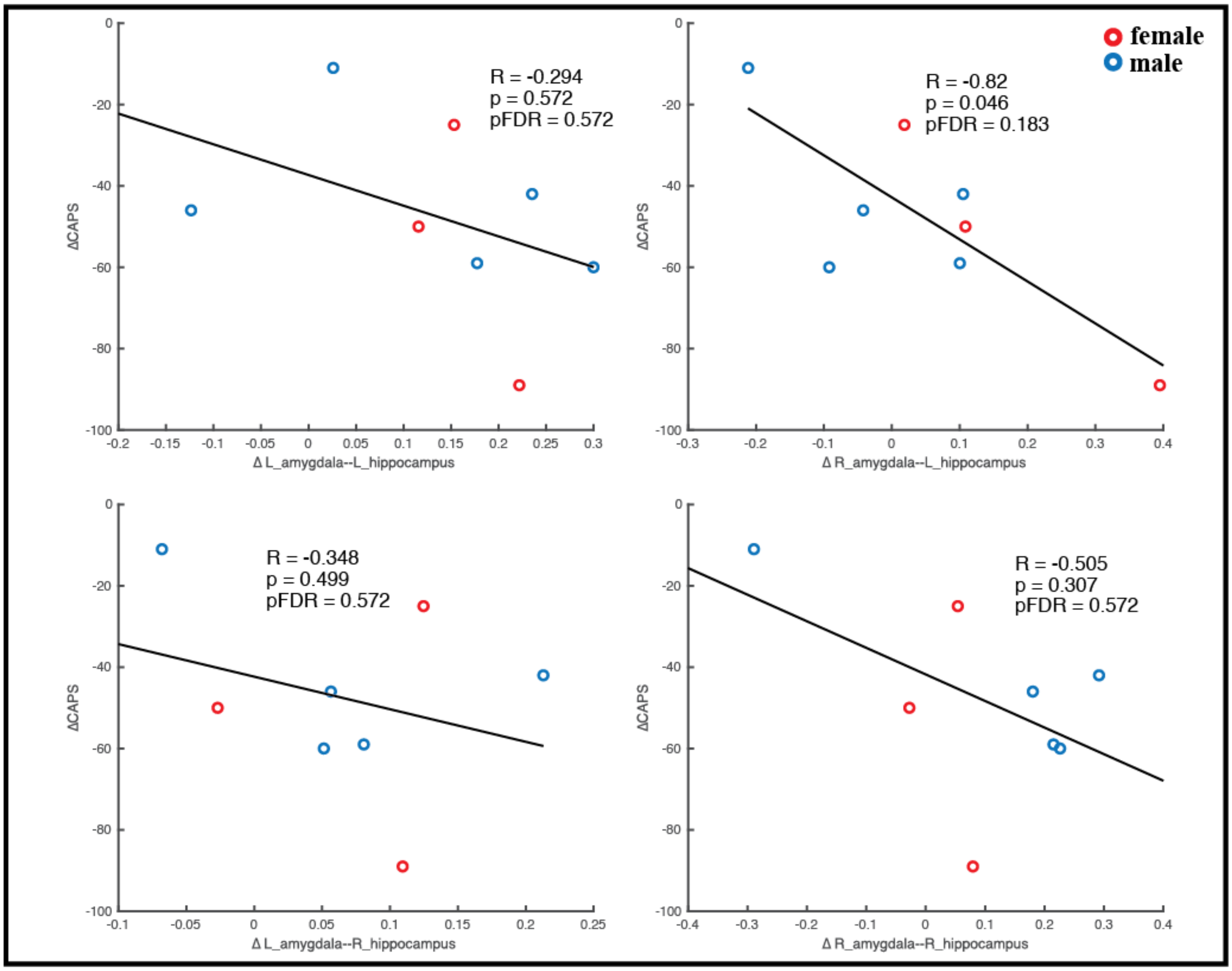


SI Figure 1: Pearson correlations (two-sided) between changes in RSFC between the R/L-amygdalae and hippocampi with changes in CAPS-IV. Changes in each metric are calculated as follow-up minus baseline. Age and mean FD difference between baseline and follow-up were included as covariates of non-interest. Red marker = female. Blue marker = male. N=8.

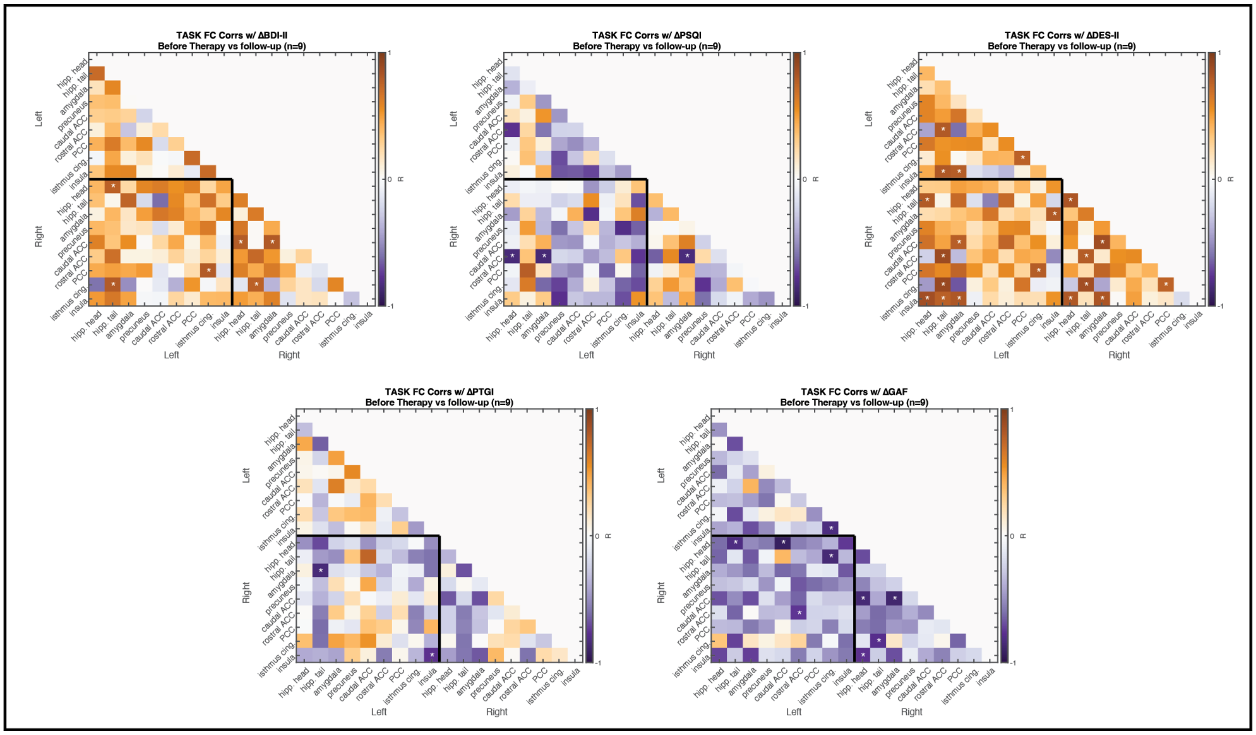


SI Figure 2: Correlations between changes in task functional connectivity and changes in secondary outcome measures. Change was calculated as follow-up values minus baseline values. Age and mean FD difference between baseline and follow-up were included as covariates of non-interest. (N=9; * two-tailed, uncorrected $p < 0.05$). No correlations were significant following multiple comparisons correction. BDI-II = Beck Depression Inventory (depressive symptoms), PSQI = Pittsburgh Sleep Quality Index (sleep quality), DES-II = Dissociative Experiences Scale (symptoms of dissociation), PTGI = Post-traumatic Growth Inventory (perceived growth following trauma), and GAF = Global Assessment of Functioning (general psychological function).

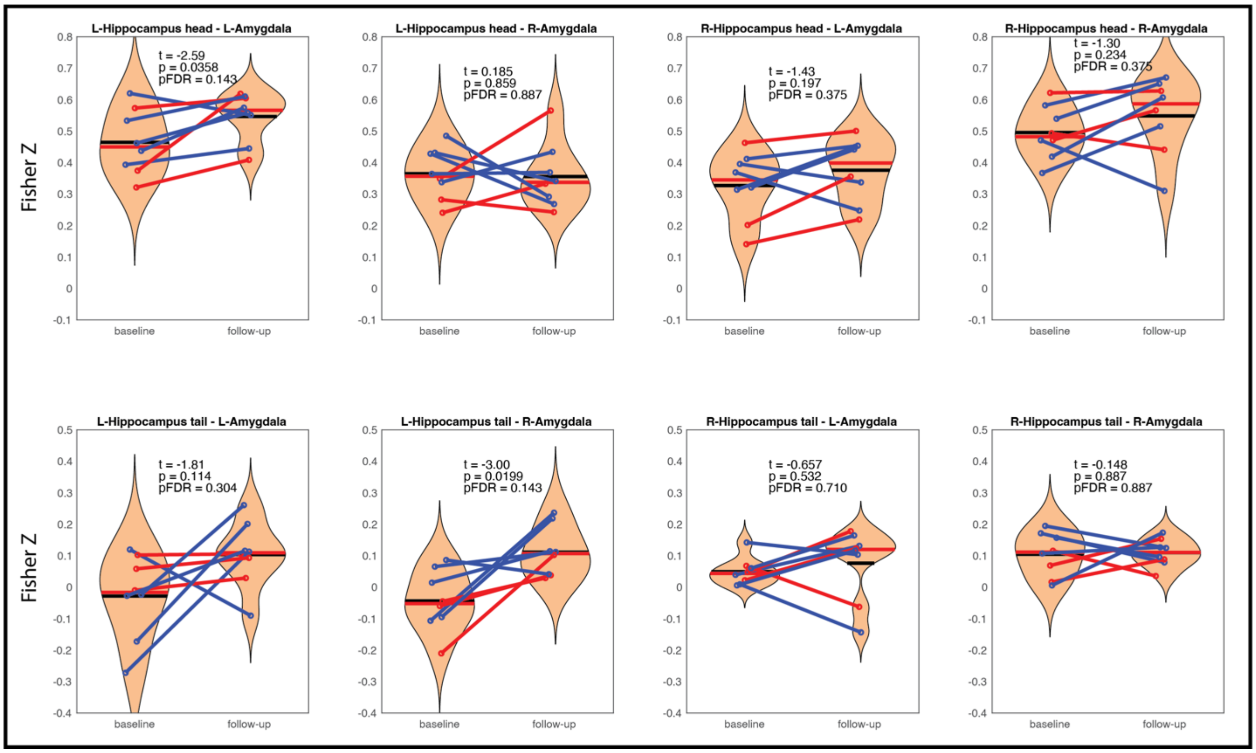


SI Figure 3: Replication of amygdala-hippocampal RSFC analyses with anterior (head) and posterior (tail) segmentation of the hippocampus. Blue marker = male. Red marker = female.

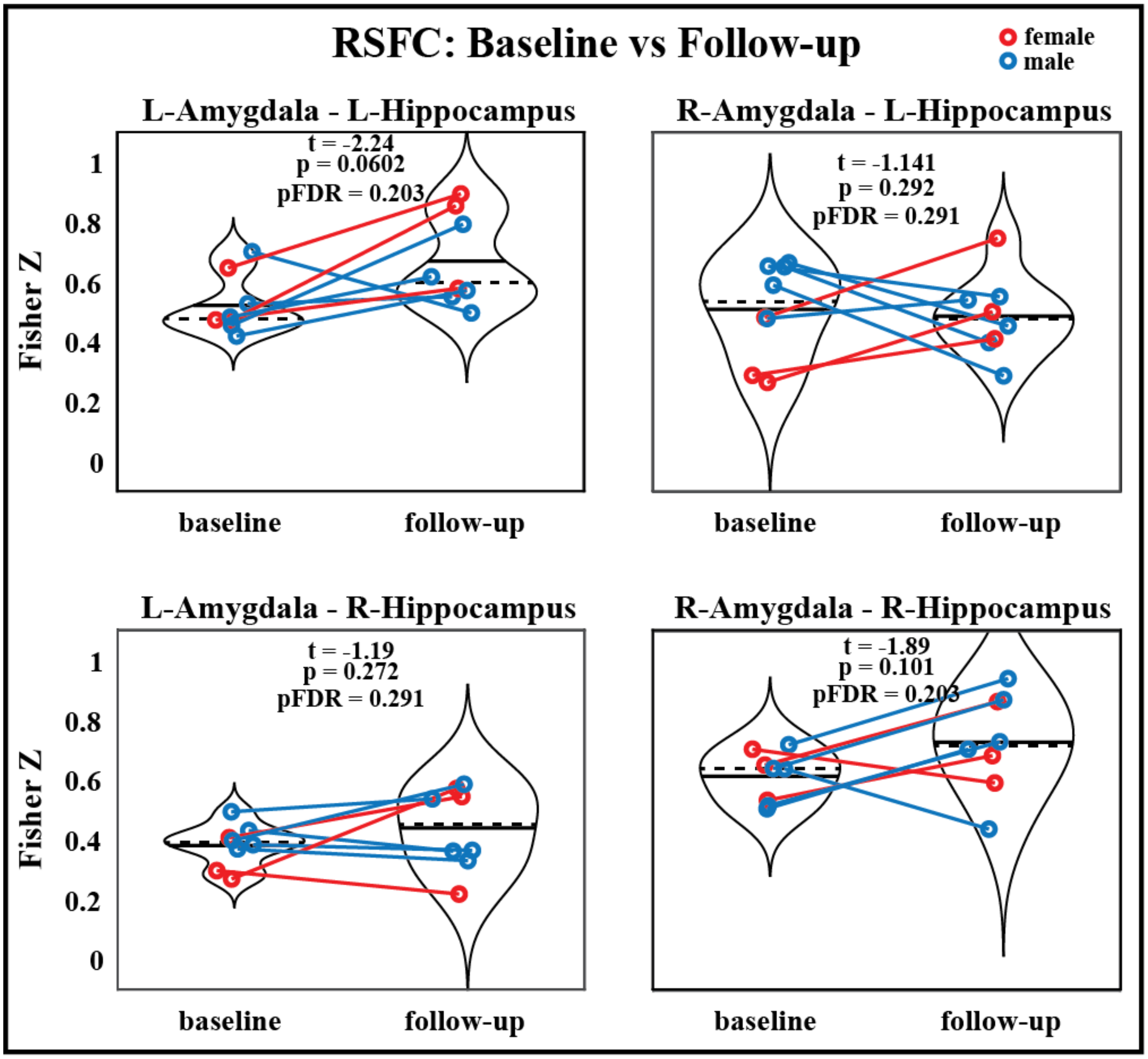


SI Figure 4: Replication of the amygdala-hippocampal RSFC analysis (main Figure 3) without the use of global signal regression. The RSFC between the L-amygdala and L-hippocampus is no longer significantly increased prior to (nor after) correction for multiple comparisons using this processing choice (p = 0.06). Blue marker = male. Red marker = female.

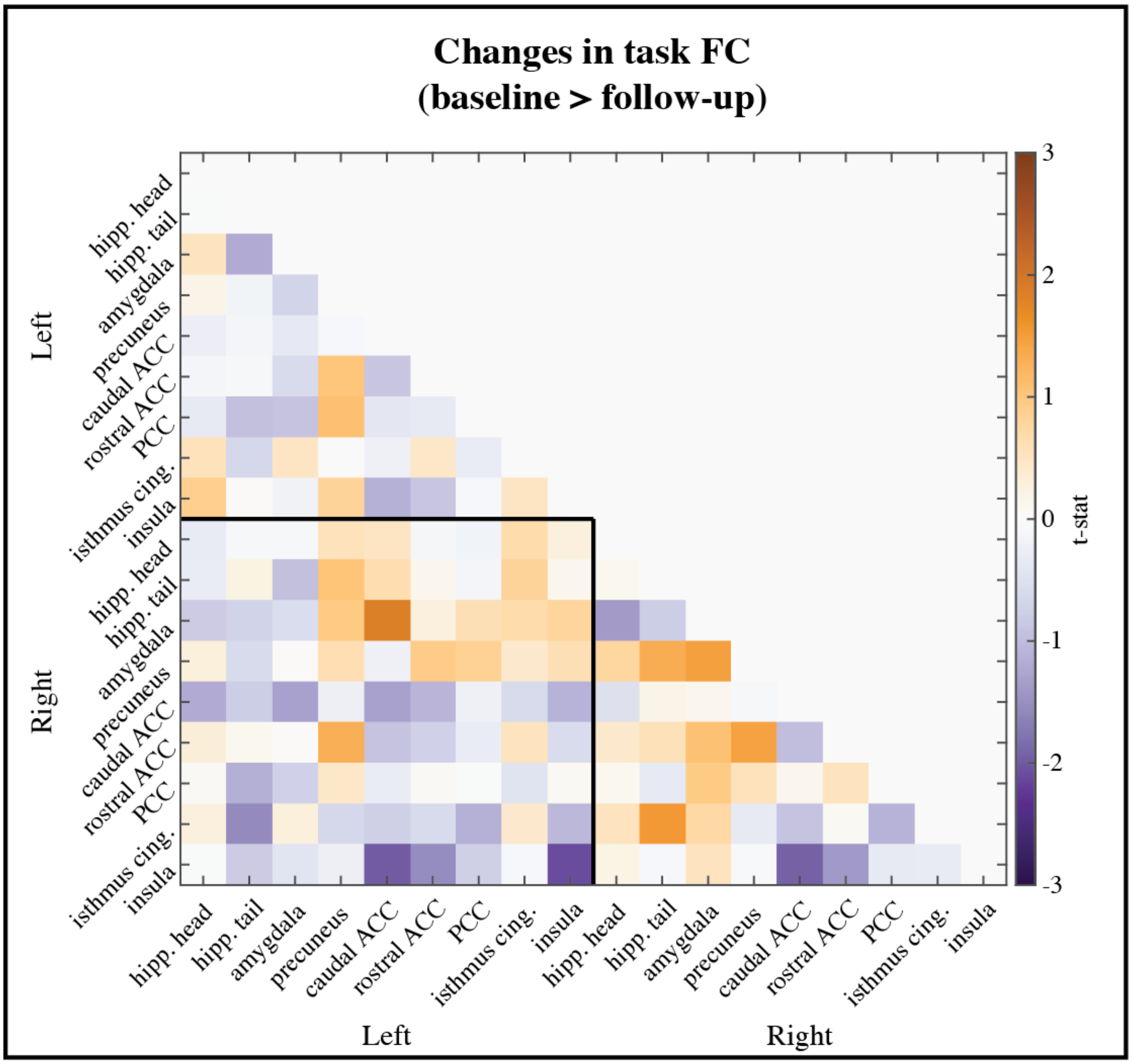


SI Figure 5: Replication of pre- to post-therapy task FC comparison (main Figure 5) without the use of global signal regression.

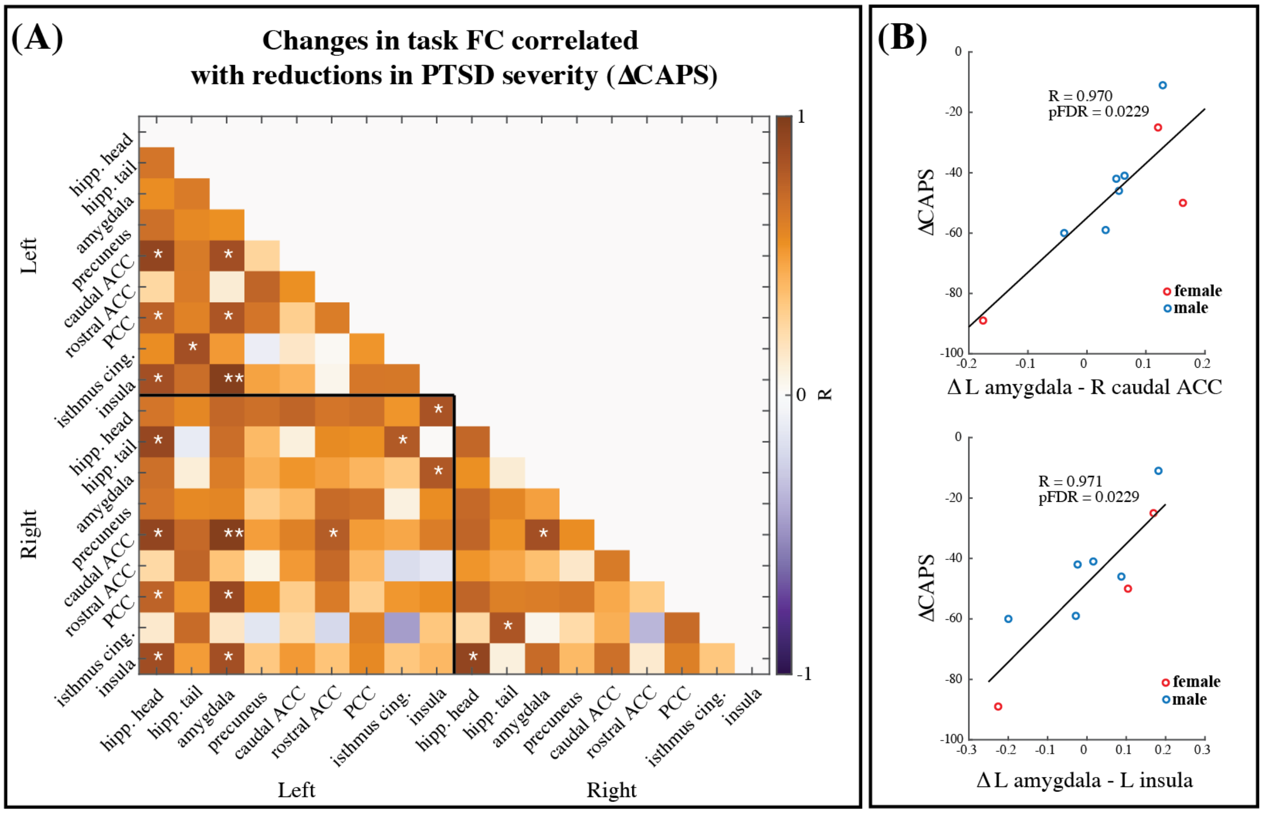


SI Figure 6: (A) Replication of the correlation between changes in task FC and reduction in CAPS scores without the use of global signal regression (N=9; * uncorrected p < 0.05, ** pFDR < 0.05). Age and mean FD difference between baseline and follow-up were included as covariates of non-interest. Many of the same functional connections that were significant with global signal regression (main Figure 6) were significant here as well. After correction for multiple comparisons, two correlations remained significant. Scatter plots of those two correlations are shown here in (B). Red marker = female. Blue marker = male.  The correlation between the left amygdala and left insula FC change and reductions in CAPS score was significant after correction for multiple comparisons in both processing choices.

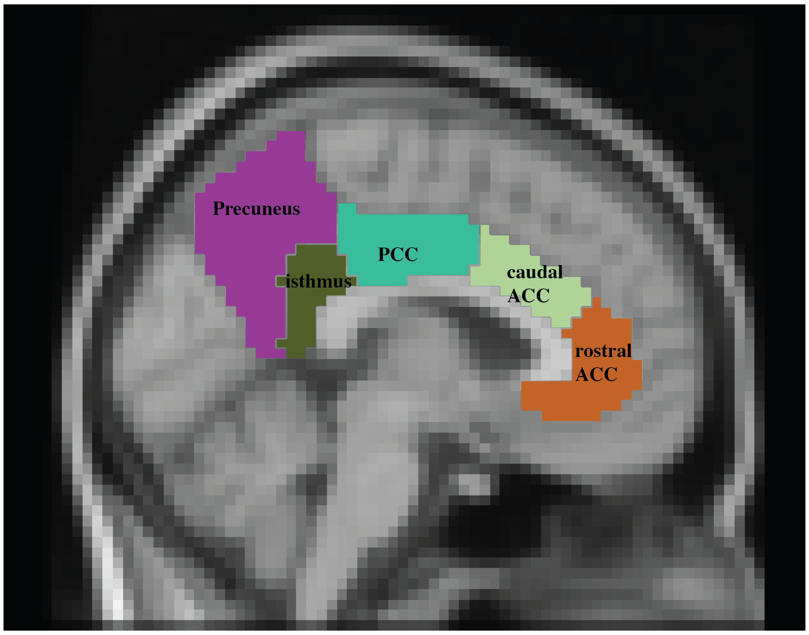


SI Figure 7: Labeling of Desikan-Killiany (DK) mid-line ROIs used in the task FC analysis, shown here for the left hemisphere. The PCC in DK begins slightly more anterior than some other atlases and thus contains portions of the cingulate that some might label as the mid-cingulate. In addition, the isthmus cingulate here overlaps with parts of the brain often considered to be the most posterior portion of the PCC.

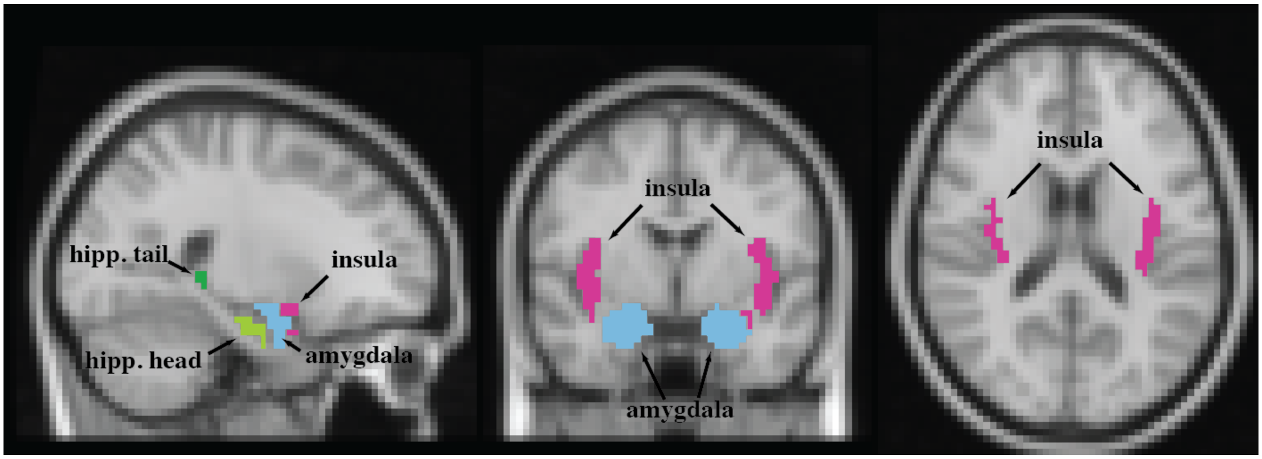


SI Figure 8: ROI definitions for the remaining DK regions used in the task FC analysis.

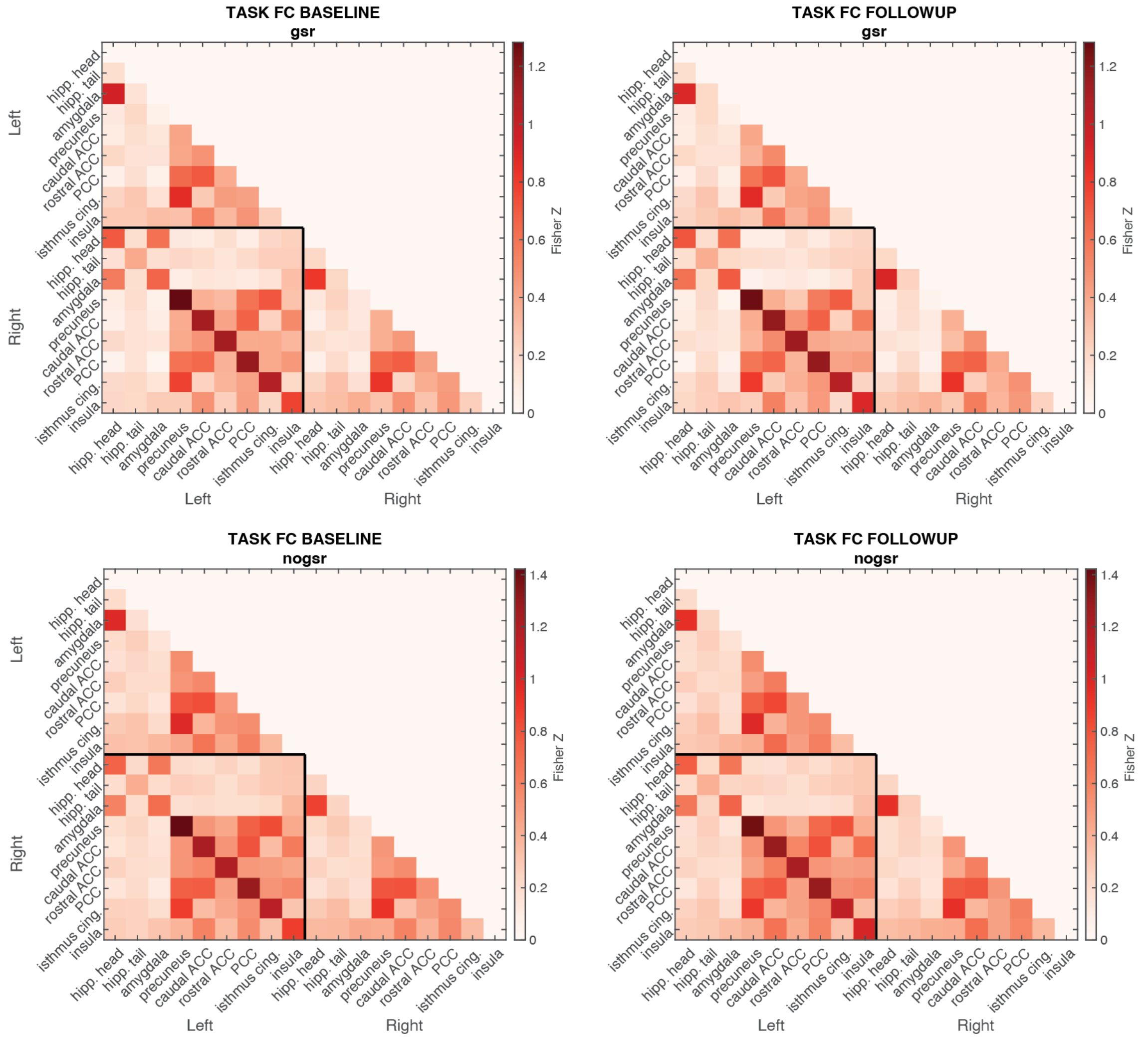


SI Figure 9: Group-level mean task FC between at each time point, with (top) and without (bottom) global signal regression (GSR).

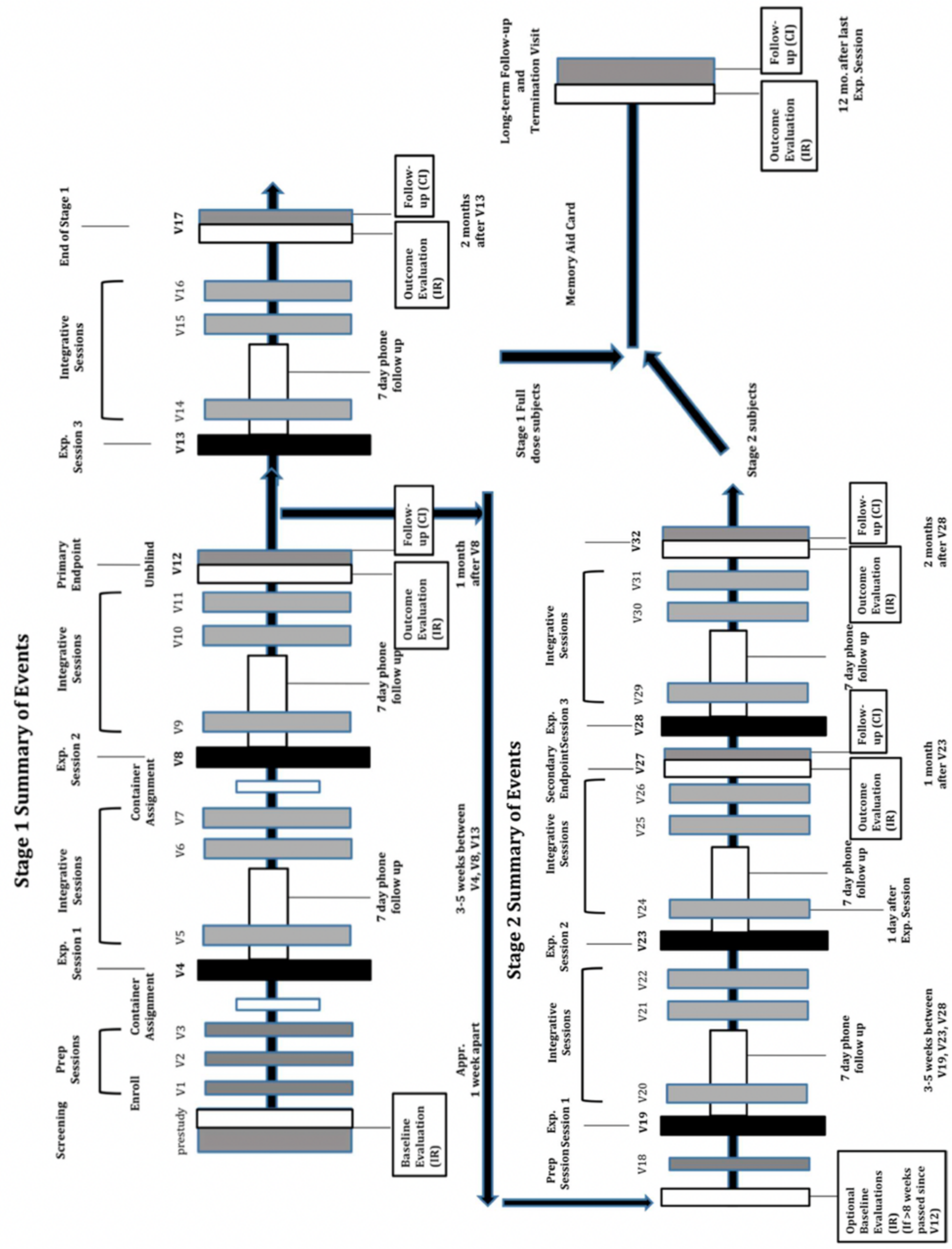

Stage 1 Summary of Events
Screening
Enroll
Prep Sessions
Container Assignment
Exp. Session 1
Integrative Sessions
Container Assignment
Exp. Session 2
Integrative Sessions
Primary Endpoint
Unblind
Exp. Session 3
Integrative Sessions
End of Stage 1
prestudy
V1
V2
V3
V4
V5
V6
V7
V8
V9
V10
V11
V12
V13
V14
V15
V16
V17
Baseline Evaluation (IR)
7 day phone follow up
Outcome Evaluation (IR)
Follow-up (CI)
1 month after V8
2 months after V13
Appr. 1 week apart
3-5 weeks between V4, V8, V13
Stage 2 Summary of Events
Prep Session
Exp. Session 1
Integrative Sessions
Exp. Session 2
Integrative Sessions
Secondary Endpoint
Exp. Session 3
Integrative Sessions
V18
V19
V20
V21
V22
V23
V24
V25
V26
V27
V28
V29
V30
V31
V32
Optional Baseline Evaluations (IR) (If >8 weeks passed since V12)
7 day phone follow up
1 day after Exp. Session
Outcome Evaluation (IR)
Follow-up (CI)
1 month after V23
2 months after V28
3-5 weeks between V19, V23, V28
Stage 1 Full dose subjects
Stage 2 subjects
Memory Aid Card
Long-term Follow-up and Termination Visit
Outcome Evaluation (IR)
Follow-up (CI)
12 mo. after last Exp. Session
